# Development and Standardization of a Classification System for Osteoradionecrosis: Implementation of a Risk-Based Model

**DOI:** 10.1101/2023.09.12.23295454

**Authors:** Erin E Watson, Katrina Hueniken, Junhyung Lee, Sophie H Huang, Amr El Maghrabi A, Wei Xu, Amy C Moreno, C. Jillian Tsai, Ezra Hahn, Andrew J McPartlin, Christopher MKL Yao, David P Goldstein, John R De Almeida, John N Waldon, Clifton David Fuller, Andrew J Hope, Salvatore L Ruggiero, Michael Glogauer, Ali A Hosni

## Abstract

**Purpose:** Osteoradionecrosis of the jaw (ORN) can manifest in varying severity. The aim of this study is to identify ORN risk factors and develop a novel classification to depict the severity of ORN.

**Methods:** Consecutive head-and-neck cancer (HNC) patients treated with curative-intent IMRT (≥45Gy) in 2011-2018 were included. Occurrence of ORN was identified from in-house prospective dental and clinical databases and charts. Multivariable logistic regression model was used to identify risk factors and stratify patients into high-risk and low-risk groups. A novel ORN classification system was developed to depict ORN severity by modifying existing systems and incorporating expert opinion. The performance of the novel system was compared to fifteen existing systems for their ability to identify and predict serious ORN event (jaw fracture or requiring jaw resection).

**Results:** ORN was identified in 219 out of 2732 (8%) consecutive HNC patients. Factors associated with high-risk of ORN were: oral-cavity or oropharyngeal primaries, received IMRT dose ≥60Gy, current/ex-smokers, and/or stage III-IV periodontal disease. The ORN rate for high-risk vs low-risk patients was 12.7% vs 3.1% (p<0.001) with an area-under-the-receiver-operating-curve (AUC) of 0.71. Existing ORN systems overclassified serious ORN events and failed to recognize maxillary ORN. A novel ORN classification system, RadORN, was proposed based on vertical extent of bone necrosis and presence/absence of exposed bone/fistula. This system detected serious ORN events in 5.7% of patients and statistically outperformed existing systems.

**Conclusion:** We identified risk factors for ORN, and proposed a novel ORN classification system based on vertical extent of bone necrosis and presence/absence of exposed bone/fistula. It outperformed existing systems in depicting the seriousness of ORN, and may facilitate clinical care and clinical trials.

## INTRODUCTION

Osteoradionecrosis (ORN) is one of the most severe radiation-induced complications for patients with head and neck cancer (HNC). ORN has been loosely defined in literature as non-healing mucosal breakdown and bone injury, occurring spontaneously or after trauma, within the head and neck (HN) radiation volume in absence of recurrent tumor on the affected site. Although these criteria are widely accepted for a clinical diagnosis, they fail to incorporate objective imaging findings^1^.

The prevalence of ORN varies widely in the literature (up to 40%) with a substantial decrease in modern era (4-8%)^2^. This overall reduction of ORN can be attributed to advances in radiotherapy technology^3,4^, and the use of a strict prophylactic dental care policy among institutions^5^. ORN may manifested in various forms reflecting its severity. The severity of ORN is depicted by various classification systems, generally defined based on: *1) clinical signs* (exposed bone, bone spicules, sequestra, infection and fistula), *2) radiographic finding* (abnormal bone pattern, radiographic sequestra and pathological fracture), and *3) treatment required and response to therapy*, either conservative (chlorhexidine rinse, antibiotics, Pentoxifylline/tocopherol - PENTOCLO, and hyperbaric oxygen - HBO) or surgical (sequestrectomy, debridement, and jaw resection). However, to the best of our knowledge, there currently is no consensus regarding the most appropriate classification system, which hinders inter-study comparison and clinical trial design. Furthermore, the most well-accepted systems overclassify serious ORN, fail to acknowledge maxillary ORN or rely on subjective findings.

Therefore, we conducted this single institution study aimed to identify risk factors for ORN in HNC patients treated with IMRT, evaluate the appropriateness of various ORN classification systems in depicting the severity of ORN, and finally to propose a novel and improved ORN classification system incorporating clinically relevant data that can be used in routine clinical care and future clinical trials.

## METHODS

### Study population and data compilation

We conducted a retrospective review of all adult HNC patients treated with curative-intent (definitive, pre-, or post-operative) IMRT ≥ 45Gy in our institution from January 1, 2011 to January 1, 2018. Patients with a previous history of HN radiation, a diagnosis of primary tracheal cancer or early-stage laryngeal cancer (T1-2 N0 M0), or edentulous patients were excluded. Ethics approval was obtained from the institutional research ethics board prior to initiation of the study.

Clinical information, and incidence and severity of ORN were retrieved from in-house prospective *Clinical* and *Dental* databases, and cross checked against individual patient dental charts to ensure the completeness and quality of the data. The *Clinical* database comprised all HNC patients who received radiotherapy where clinical information including baseline characteristics, staging, treatment, oncologic outcomes, and toxicity data (including ORN) were recorded at point-of-care^6^. *Dental* database comprised all HNC patients who were referred to designated dental clinic prior to receiving radiotherapy and continuous dental care after completion of treatment. Their dental care information was registered prospectively and updated after each dental visit. Final extracted patient parameters from the *Dental* and *Clinical* databases included patients’ demographics (age, gender, smoking, postal code as surrogate for gross income), dental insurance status (private insurance, self-pay or public insurance), cancer characteristics (primary site, staging and treatment) and dental characteristics (DMFS160)^7^, number of teeth at baseline, number of teeth removed prior to and remaining after RT, Periodontal status^8^). Pre-IMRT periodontal condition (PC) was classified by universal multi-dimensional staging system based on number of teeth and amount of bone loss (stages 0 to IV)^8^.

#### Assessment and Classification of ORN severity

All data related to ORN was independently reviewed by two reviewers. Each individual visit for all patients with ORN was reviewed and ORN events were retrospectively classified according to fifteen different ORN classification systems [**Supplement Table 1**]: Coffin^9^, Marx^10^, Morton and Simpson^11^, Radiation Therapy Oncology Group/European Organization for Research and Treatment of Cancer (RTOG/EORTC)^12^, Glanzmann and Gratz^13^, Clayman^14^, Late Effects Normal Tissue Task Force Subjective, Objective, Management, Analytic scales (LENT/SOMA)^15^, Store and Boysen^16^, Schwartz and Kagan^17^, Notani^18^, Tsai^19^, Lyons^20^, Common Terminology Criteria for Adverse Events, CTCAE^21^, Caparrotti ^22^ and Shaw^23^. These ORN classification systems were widely used in dental and oncology literature and practice.

Distributions of classifications of ORN for each system were summarized across all visits. Proportions of visits *<u>missing</u>* sufficient data to assign a classification in each system were summarized, and presented both unweighted (all visits weighted equally) and weighted (mean proportion of missing visits was first calculated for each patient, then aggregated so that each patient contributed equal weight). Proportions of patients with at least one classification in the *<u>most severe category</u>* for each system were summarized.

#### Statistical Analysis

### Identifying ORN Risk Factors and Stratifying Risk Groups

Univariable analysis (UVA) with logistic regression was conducted to assess the association of the following clinical factors with ORN: primary tumor location, stage, gender, smoking status, RT dose, concurrent chemotherapy, surgical management, PC, age at diagnosis, and smoking pack/years. Variables with p<0.1 in UVA were included in a multivariable ORN risk-score model which classified patients into high and low risk groups [13]. Total risk scores were computed by adding the resulting model coefficients on the log scale (termed “risk score coefficients”). An optimal cutpoint for classifying patients as high-risk vs low-risk was identified by maximizing Youden’s J^24^. To measure predictive performance of the binary high-risk vs low-risk scores, area under the receiver-operating curve (AUC) was calculated from repeated 10-fold cross-validation with 10 iterations. All tests were two-sided, and used p<0.05 to define statistical significance.

### Evaluating the Performance of Various Classification Systems

The performance of ORN classification systems were evaluated for “*<u>separability</u>*”, defined here as the ability of each system to discriminate between patients who would go on to experience a “serious ORN event” (defined by jaw fracture or ORN requiring jaw resection) versus those who would not. First, Kaplan-Meier curves and logrank tests were used to compare time from ORN detection to serious ORN event, stratified by stage/grade at the time of ORN detection. Second, univariable Cox proportional hazards regression models were fit to evaluate time to serious ORN event from ORN stage/grade at any visit post-detection of ORN, treating ORN stage as a time-varying covariate. Finally, predictive performance of each classification system was evaluated via model concordance and time-dependent AUC at several clinically-relevant timepoints, using the methods described by Bansal and Heagerty^25^. Time-dependent AUC measures classification accuracy between patients with a serious ORN event vs. no events at a given landmark time. Separability analysis was not performed on systems for which >50% of all visits were missing/non-evaluable.

AUC is derived from measures of sensitivity and specificity of each ORN classification system. AUCs were computed using two different methods of measuring sensitivity^25,26^. 1) “Incident sensitivity” was defined as the probability of high stage at a given time t, among patients who had jaw fracture or surgery at time t. 2) “Cumulative sensitivity” was defined as the probability of high stage at time t, among patients who would go on to experience jaw fracture or surgery within the interval (t, t + one year]. Both methods used “dynamic specificity”, the probability of low stage at time t among patients who were event-free at time t. Incident and cumulative sensitivity and dynamic specificity were estimated using a nearest-neighbors estimator. AUCs for incident sensitivity/dynamic specificity (AUC I/D) were calculated at 1-, 2-, and 3-years after ORN detection. AUCs for cumulative sensitivity/dynamic specificity (AUC C/D) were calculated at one year (predicting jaw fracture/surgery between 1-2 years) and two years (predicting jaw fracture/surgery between 2-3 years). Finally, a c-index was computed for all systems to measure model concordance, derived from a weighted average of AUC I/D values across the entire follow-up period^25^. Higher c-index indicates better ability for a grading system to discriminate between patients who will have adverse outcomes versus none. Confidence intervals for all AUCs and c-indices were obtained via bootstrapping with 1000 iterations.

### Proposing a Novel ORN Classification System

The existing ORN classification systems that were used to inform the novel classification system were those identified as high-performing on our analysis based on proportion of missingness (and most-severe stage or grade), and separability (i.e., time to serious negative ORN outcomes assessed by Kaplan-Meier curves from first ORN detection, time-to-event cox regression treating ORN stage as a time-varying covariate, and time-dependent AUC). The components of these systems were further analyzed in light of their statistical performance, with extraction of specific clinically relevant characteristics from the following ORN staging systems: 1) The Notani ^18^ staging system: recognition that early stage ORN is confined to alveolar bone based on imaging; 2) Shaw system (modified Notani)^23^: Addition of Minor Bony Spicules (MBS); 3) Store – recognition of radiographic ORN with intact mucosa^16^; 4) CTCAE^21^ – aligning objective grading criteria with treatment recommendations, and; 5) Multiple systems that recognize fracture and orocutaneous fistula as representing the most severe form of ORN ^11–13,15–20,22,23,27^. In addition, The American Association of Oral and Maxillofacial surgeons (AAOMS) staging system for MRONJ^28,29^ was utilized, in particular the recognition of ORN detectable radiographically but not clinically ^14^; stage I disease confined to alveolar bone; recognition of early changes suggestive of disease (stage 0) and inclusion of the maxilla. Finally, the final staging system also considered the results of the Delphi Study on staging of ORN currently underway at MD Anderson. A subsequent statistical analysis was undertaken to evaluate the performance of the proposed system.

## RESULTS

### Clinical characteristics

A total of 2732 HNC patients were included. Median age was 61 years; 30% (n=807) were current/ex-smokers; 53% (n=1410) had stage III-IV PC; 18% (n=490) were oral cavity cancer (OCC), and 35% (n= 969) were oropharyngeal cancer (OPC). The median IMRT dose was 70 Gy/35 fractions; 43% (n=1170) received concurrent chemotherapy; and 37% (n=1006) underwent surgery (**Table 1**). A total of 219 patients (8%) developed ORN.

**Table 1:** Patient, Tumor and treatment characteristics.

| **Covariate** | **Full Sample**<br>**(n=2732)** | **ORN (No)**<br>**(n=2508)** | **ORN (Yes)**<br>**(n=224)** | **p-value** |
| --- | --- | --- | --- | --- |
| **Insurance Type** |  |  |  | **<0.001** |
| No Insurance | 1160 (42) | 1076 (43) | 84 (38) |  |
| Private | 1373 (50) | 1264 (50) | 109 (49) |  |
| Public | 199 (7) | 168 (7) | 31 (14) |  |
| **Teeth-preRT [median (range)]** | 25 (1-32) | 25 (1-32) | 25 (2-32) | 0.60 |
| **Teeth-PostRT [median**<br>**(range)]** | 24 (0-32) | 24 (0-32) | 23 (0-32) | 0.35 |
| **DMFS [median (range)]** | 71 (0-160) | 71 (0-160) | 72 (5,160) | 0.86 |
| **Smoking** |  |  |  | **<0.001** |
| No | 1925 (70) | 1791 (71) | 134 (60) |  |
| Yes | 807 (30) | 717 (29) | 90 (40) |  |
| **Chemotherapy** |  |  |  | 0.28 |
| No | 1563 (57) | 1443 (58) | 120 (54) |  |
| Yes | 1169 (43) | 1065 (42) | 104 (46) |  |
| **Surgery** |  |  |  | 1 |
| No | 1727 (63) | 1585 (63) | 142 (63) |  |
| Yes | 1005 (37) | 923 (37) | 82 (37) |  |
| **Age at Dx [median (range)]** | 61.0 (18.3-96.7) | 61.3 (18.3-96.7) | 59.0 (27.7-88.9) | 0.082 |
| **Gender** |  |  |  | 0.82 |
| Female | 732 (27) | 670 (27) | 62 (28) |  |
| Male | 2000 (73) | 1838 (73) | 162 (72) |  |
| **N category** |  |  |  | 0.084 |
| N0 | 790 (29) | 722 (29) | 68 (30) |  |
| N1 | 351 (13) | 324 (13) | 27 (12) |  |
| N2 | 1405 (52) | 1282 (52) | 123 (55) |  |
| N3 | 156 (6) | 151 (6) | 5 (2) |  |
| Missing | **30** | **29** | **1** |  |
| **Disease Site** |  |  |  | **<0.001** |
| Other | 1074 (39) | 1037 (41) | 37 (17) |  |
| Larynx | 199 (7) | 197 (8) | 2 (1) |  |
| Lip & Oral Cavity | 490 (18) | 415 (17) | 75 (33) |  |
| Oropharynx | 969 (35) | 859 (34) | 110 (49) |  |
| **Teeth Removed** |  |  |  | 0.2 |
| Mean (sd) | 1.3 (3.0) | 1.3 (2.9) | 1.7 (3.6) |  |
| Median (Min,Max) | 0 (0,28) | 0 (0,26) | 0 (0,28) |  |
| Missing | **1** | **1** | **0** |  |
| **Proportion Teeth Removed** |  |  |  | 0.23 |
| Mean (sd) | 0.1 (0.2) | 0.1 (0.2) | 0.1 (0.2) |  |
| Median (Min,Max) | 0 (0,1) | 0 (0,1) | 0 (0,1) |  |
| Missing | **1** | **1** | **0** |  |
| **Smoking PY [median (range)]** | 26 (1-89) | 21 (1-89) | 35 (1-86) | **<0.001** |
| **PC** |  |  |  | **0.012** |
| I | 270 (10) | 252 (10) | 18 (8) |  |
| II | 504 (19) | 466 (19) | 38 (17) |  |
| III | 747 (28) | 673 (27) | 74 (33) |  |
| IV | 663 (25) | 596 (24) | 67 (30) |  |
| O | 489 (18) | 463 (19) | 26 (12) |  |
| Missing | **59** | **58** | **1** |  |
| **Pre-RT Extraction** |  |  |  | 0.3 |
| No | 1776 (65) | 1638 (65) | 138 (62) |  |
| Yes | 956 (35) | 870 (35) | 86 (38) |  |
| **RT Dose** [median (range)] | 70.0 (2.0-76.7) | 70.0 (2.0-76.7) | 70 (50-70) | 0.16 |
| Abbreviations: DMFS: Decayed, missing, filled surfaces according to DMFS160; Pre-RT: before commencement of radiotherapy; post-RT: after completion of radiotherapy; PY: pack-years; PC: Periodontal Condition |  |  |  |  |

### ORN Risk Factors and Risk Groups

MVA (**Table 2)** identified the following ORN risk factors: current/ex-smoker (OR:1.48, 95% CI: 1.1-1.99; p=0.01; RSC=0.4), stage III-IV PC (OR:1.55, 95%CI: 1.15-2.08; p<0.01; RSC=0.4), primary OCC (OR:5.66, 95% CI: 3.74-8.55; p<0.001; RSC=1.7), primary OPC (OR:4.05, 95% CI: 2.75-5.96; p<0.001; RSC=1.4), and IMRT dose prescription ≥60Gy (OR:3.59, 95% CI: 1.12-11.51; p=0.03; RSC=1.3). Patients were classified into ORN risk groups according to the sum of their risk score coefficients. High-risk group was defined by the sum of risk scores ≥3.5, while low-risk group was defined by a sum under 3.5. Applying the generated model, the high-risk group included OCC or OPC patients who received IMRT≥60Gy and current/ex-smoker and/or stage III-IV PC. The overall rate of ORN for high-risk patients was 12.7% (95% CI 11-14.5%) as compared to 3.1% (95% CI: 2.1-4%) in the low-risk group (p<0.001). Model Performance from 10-fold cross-validation showed an AUC of 0.71 (95% CI: 0.61-0.81).

**Table 2:** Multivariable analysis of risk factors for osteoradionecrosis.

|  | Odds Ratio<br>(95%CI) | p-value | Global p-value | N |
| --- | --- | --- | --- | --- |
| **Smoking** |  | **0.010** |  | 2673 |
| No | Reference |  |  | 1896 |
| Yes | 1.48 (1.10,<br>1.99) |  |  | 777 |
| **Primary Site** |  |  | **<0.001** | 2673 |
| Larynx/Other | Reference |  |  | 1246 |
| Lip/Oral Cavity | 5.66 (3.74,<br>8.55) | **<0.001** |  | 476 |
| Oropharynx | 4.05 (2.75,<br>5.96) | **<0.001** |  | 951 |
| **Dose** |  | **0.03** |  | 2673 |
| <60 | Reference |  |  | 144 |
| >=60 | 3.59 (1.12,<br>11.51) |  |  | 2529 |
| **Periodontal**<br>**Classification** |  | **0.004** |  | 2673 |
| 0-2 (Mild) | Reference |  |  | 1263 |
| 3-4 (Moderate-severe) | 1.55 (1.15,<br>2.08) |  |  | 1410 |

### Comparison of Various ORN Classification systems

Missingness (the average proportion of visits where an ORN stage could not be generated per patient by each grading system) is summarized in **Table 3**. The following ORN grading systems showed less than 25% missingness: Clayman (5.4%), CTCAE (20.6%), Notani (20.9%), RTOG/EORTC (19.3%) and Store (7.5%) systems. The proportion of patients with at least one most-severe stage is summarized in **Table 3**. The following ORN classification systems detected less than 5% of patients as having the most severe stage or grade of ORN: RTOG/EORTC (1.1%), SOMA (4.7%), Store (4.9%) and Schwartz (2.5%) systems.

**Table 3:**
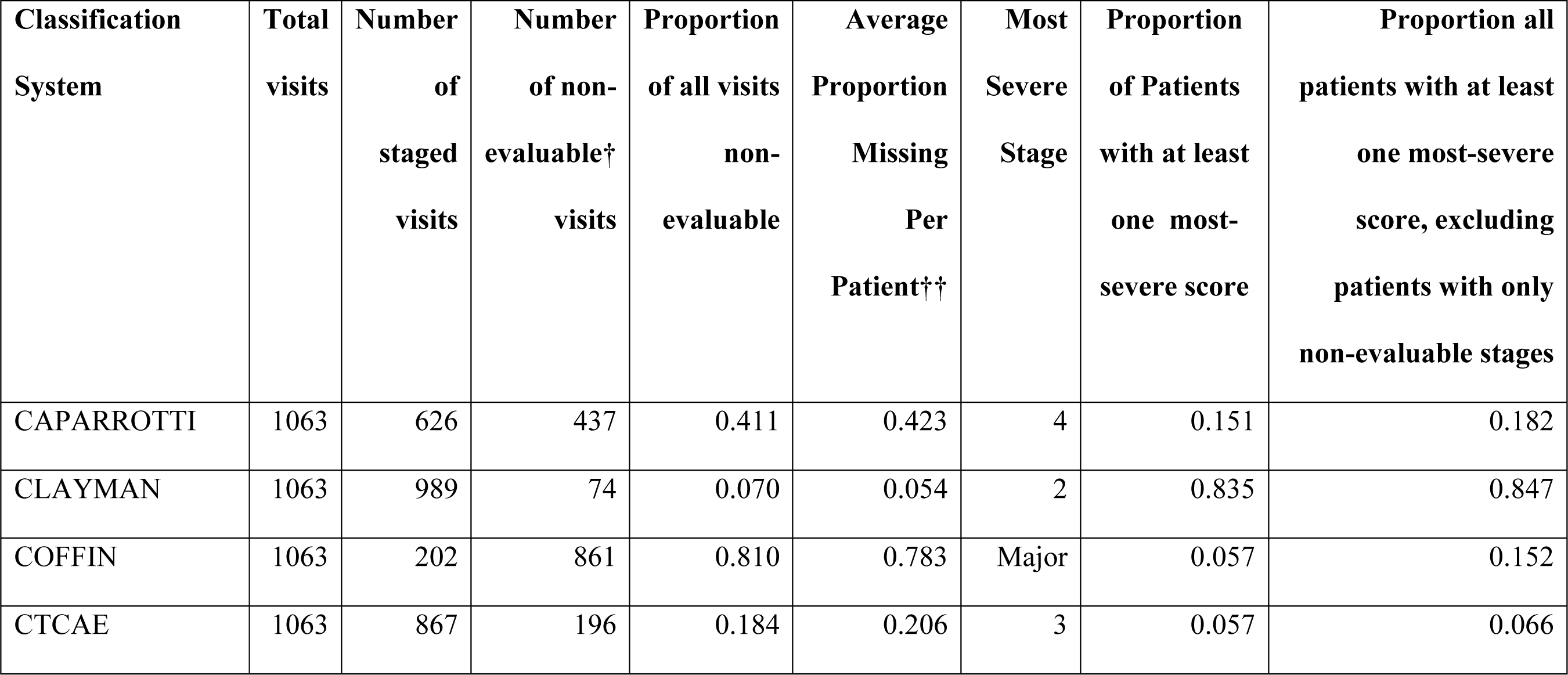
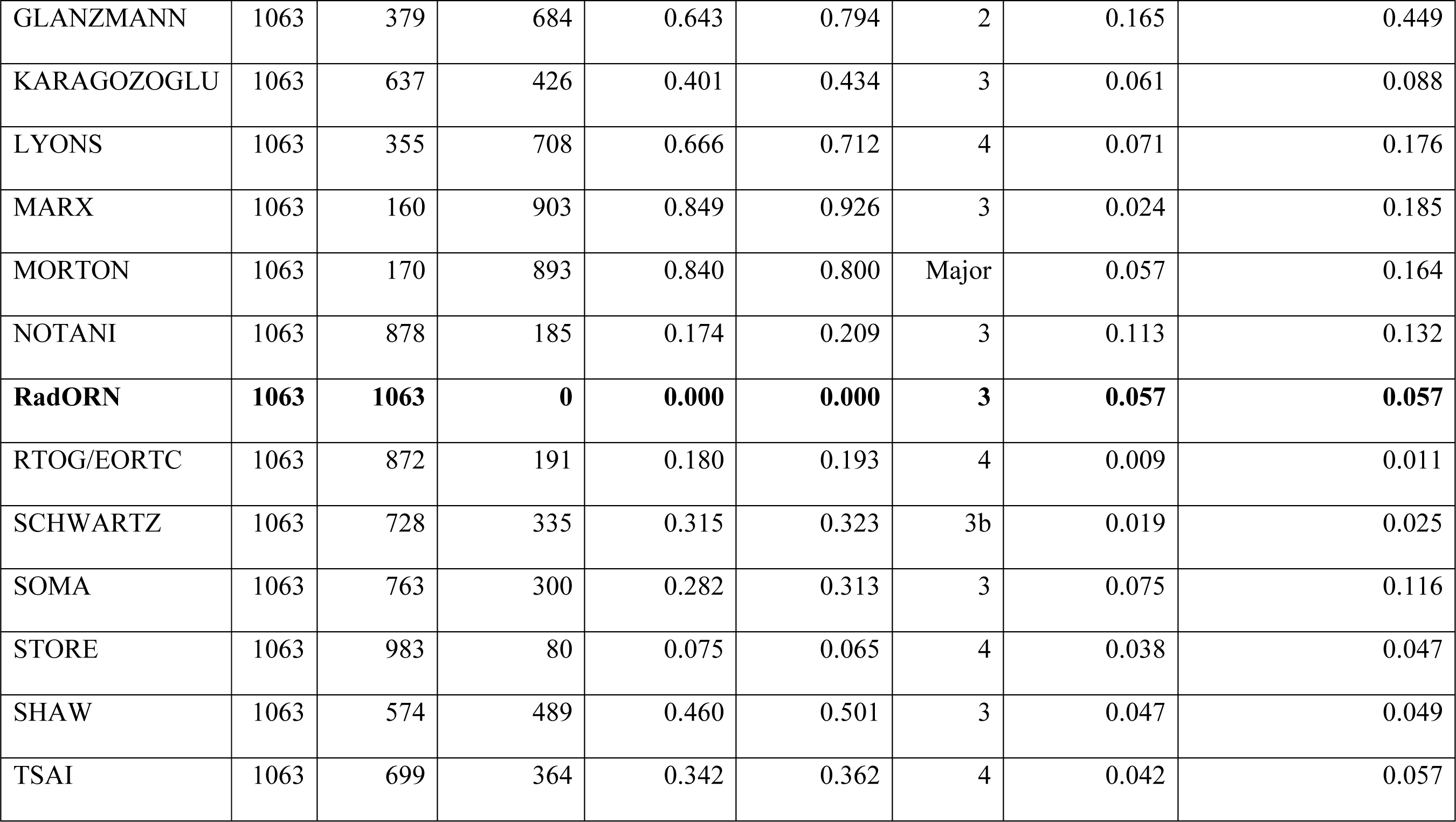
Count and proportion of visits missing staging information in each system and proportion of all patients with at least one visit with most-severe stage.

| **Classification**<br>**System** | **Total**<br>**visits** | **Number**<br>**of**<br>**staged**<br>**visits** | **Number**<br>**of non-**<br>**evaluable†**<br>**visits** | **Proportion**<br>**of all visits**<br>**non-**<br>**evaluable** | **Average**<br>**Proportion**<br>**Missing**<br>**Per**<br>**Patient††** | **Most**<br>**Severe**<br>**Stage** | **Proportion**<br>**of Patients**<br>**with at least**<br>**one most-**<br>**severe score** | **Proportion all**<br>**patients with at least**<br>**one most-severe**<br>**score, excluding**<br>**patients with only**<br>**non-evaluable stages** |
| --- | --- | --- | --- | --- | --- | --- | --- | --- |
| CAPARROTTI | 1063 | 626 | 437 | 0.411 | 0.423 | 4 | 0.151 | 0.182 |
| CLAYMAN | 1063 | 989 | 74 | 0.070 | 0.054 | 2 | 0.835 | 0.847 |
| COFFIN | 1063 | 202 | 861 | 0.810 | 0.783 | Major | 0.057 | 0.152 |
| CTCAE | 1063 | 867 | 196 | 0.184 | 0.206 | 3 | 0.057 | 0.066 |
| GLANZMANN | 1063 | 379 | 684 | 0.643 | 0.794 | 2 | 0.165 | 0.449 |
| KARAGOZOGLU | 1063 | 637 | 426 | 0.401 | 0.434 | 3 | 0.061 | 0.088 |
| LYONS | 1063 | 355 | 708 | 0.666 | 0.712 | 4 | 0.071 | 0.176 |
| MARX | 1063 | 160 | 903 | 0.849 | 0.926 | 3 | 0.024 | 0.185 |
| MORTON | 1063 | 170 | 893 | 0.840 | 0.800 | Major | 0.057 | 0.164 |
| NOTANI | 1063 | 878 | 185 | 0.174 | 0.209 | 3 | 0.113 | 0.132 |
| **RadORN** | **1063** | **1063** | **0** | **0.000** | **0.000** | **3** | **0.057** | **0.057** |
| RTOG/EORTC | 1063 | 872 | 191 | 0.180 | 0.193 | 4 | 0.009 | 0.011 |
| SCHWARTZ | 1063 | 728 | 335 | 0.315 | 0.323 | 3b | 0.019 | 0.025 |
| SOMA | 1063 | 763 | 300 | 0.282 | 0.313 | 3 | 0.075 | 0.116 |
| STORE | 1063 | 983 | 80 | 0.075 | 0.065 | 4 | 0.038 | 0.047 |
| SHAW | 1063 | 574 | 489 | 0.460 | 0.501 | 3 | 0.047 | 0.049 |
| TSAI | 1063 | 699 | 364 | 0.342 | 0.362 | 4 | 0.042 | 0.057 |

Separability of different classification systems is illustrated in **Figure 1**. A total of 168 patients had sufficient ORN staging and follow-up data for separability analysis, 32 patients had a serious ORN event (i.e. jaw fracture or surgery). For the Notani, Shaw (stages 2-3 grouped), RTOG/EORTC (grades 2-4 grouped), Store (grades 2-3 grouped), Schwartz and Karagozoglu (grades 1-3 grouped) systems, patients with a more severe stage or grade at time of ORN detection were more likely to develop a serious ORN event (fracture or surgery) at an earlier timepoint than patients with each successively less severe grade. On Univariable Cox regression (**Table 4**), diagnosis with most severe stages of the Notani (stage 2-3), Shaw (stage 2-3) and RTOG/EORTC (grades 2-4) systems significantly associated with risk of serious ORN event (fracture or surgery) when compared to patients only diagnosed with the least severe stage/grade(s).

**Figure 1:**
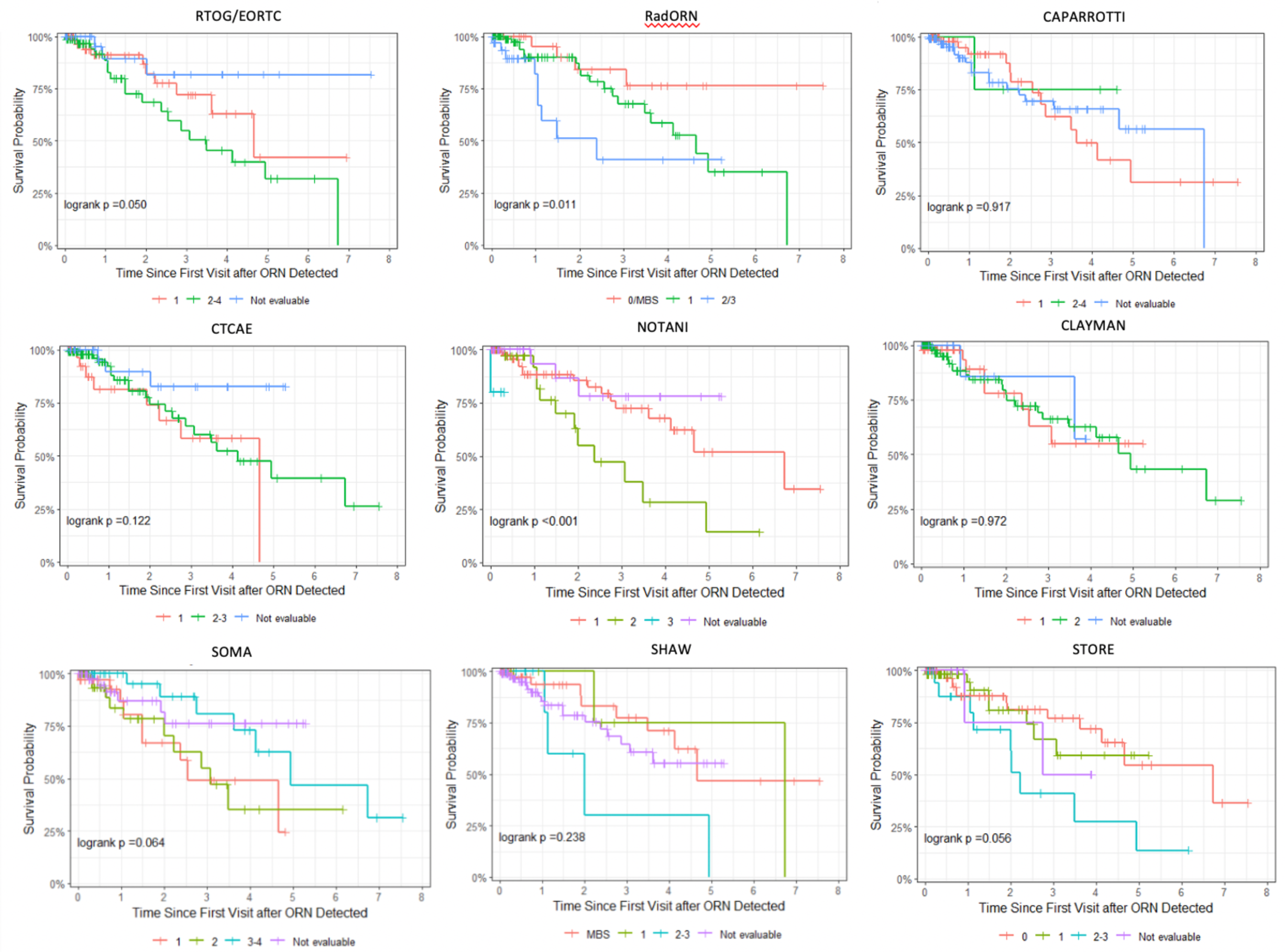
Kaplan-Meier curves for time-to-jaw-fracture or jaw surgery, stratified by stage or grade at baseline (time of ORN detection) within each classification system.

**Table 4:**
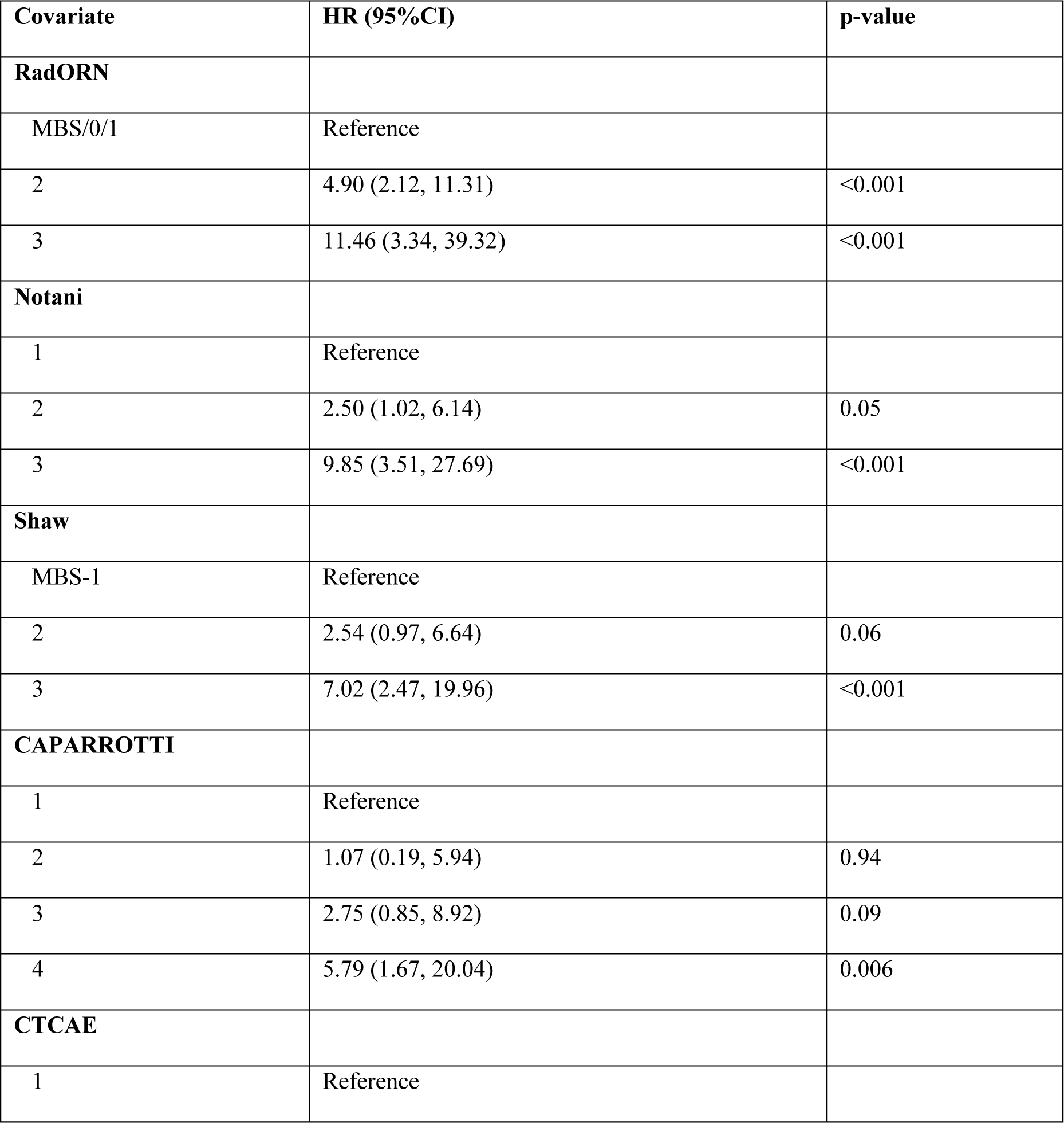
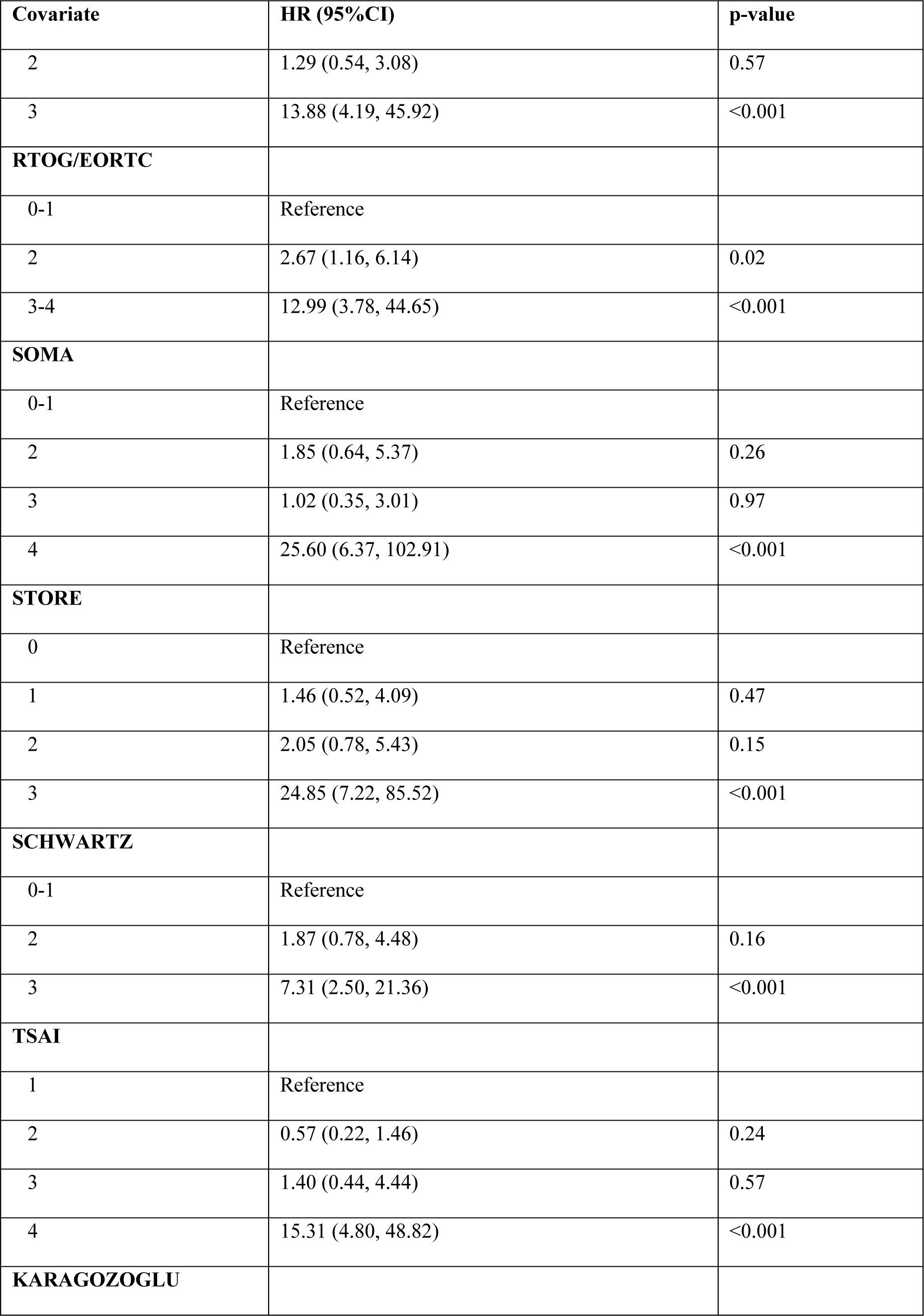
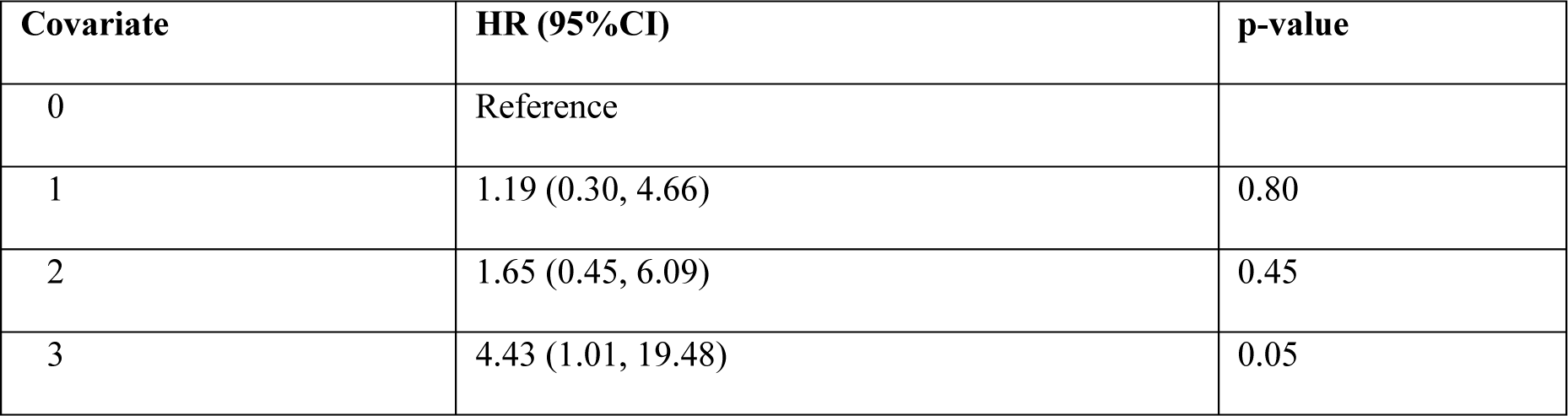
Univariable Cox time-to-event regression models: Time to jaw fracture or surgery by stage at all visits prior to fracture, as a time-varying covariate.

| Covariate | HR (95%CI) | p-value |
| --- | --- | --- |
| **RadORN** |  |  |
| MBS/0/1 | Reference |  |
| 2 | 4.90 (2.12, 11.31) | <0.001 |
| 3 | 11.46 (3.34, 39.32) | <0.001 |
| **Notani** |  |  |
| 1 | Reference |  |
| 2 | 2.50 (1.02, 6.14) | 0.05 |
| 3 | 9.85 (3.51, 27.69) | <0.001 |
| **Shaw** |  |  |
| MBS-1 | Reference |  |
| 2 | 2.54 (0.97, 6.64) | 0.06 |
| 3 | 7.02 (2.47, 19.96) | <0.001 |
| **CAPARROTTI** |  |  |
| 1 | Reference |  |
| 2 | 1.07 (0.19, 5.94) | 0.94 |
| 3 | 2.75 (0.85, 8.92) | 0.09 |
| 4 | 5.79 (1.67, 20.04) | 0.006 |
| **CTCAE** |  |  |
| 1 | Reference |  |

| **Covariate** | **HR (95%CI)** | **p-value** |
| --- | --- | --- |
| 2 | 1.29 (0.54, 3.08) | 0.57 |
| 3 | 13.88 (4.19, 45.92) | <0.001 |
| **RTOG/EORTC** |  |  |
| 0-1 | Reference |  |
| 2 | 2.67 (1.16, 6.14) | 0.02 |
| 3-4 | 12.99 (3.78, 44.65) | <0.001 |
| **SOMA** |  |  |
| 0-1 | Reference |  |
| 2 | 1.85 (0.64, 5.37) | 0.26 |
| 3 | 1.02 (0.35, 3.01) | 0.97 |
| 4 | 25.60 (6.37, 102.91) | <0.001 |
| **STORE** |  |  |
| 0 | Reference |  |
| 1 | 1.46 (0.52, 4.09) | 0.47 |
| 2 | 2.05 (0.78, 5.43) | 0.15 |
| 3 | 24.85 (7.22, 85.52) | <0.001 |
| **SCHWARTZ** |  |  |
| 0-1 | Reference |  |
| 2 | 1.87 (0.78, 4.48) | 0.16 |
| 3 | 7.31 (2.50, 21.36) | <0.001 |
| **TSAI** |  |  |
| 1 | Reference |  |
| 2 | 0.57 (0.22, 1.46) | 0.24 |
| 3 | 1.40 (0.44, 4.44) | 0.57 |
| 4 | 15.31 (4.80, 48.82) | <0.001 |
| **KARAGOZOGLU** |  |  |
| 0 | Reference |  |
| 1 | 1.19 (0.30, 4.66) | 0.80 |
| 2 | 1.65 (0.45, 6.09) | 0.45 |
| 3 | 4.43 (1.01, 19.48) | 0.05 |

### Development and evaluation of novel ORN staging and grading system

The proposed ORN classification system categorizes ORN of the jaw (ICD FB81.5) as radiographic lytic or mixed sclerotic lesions of bone and/or exposed bone or bone probed through a periodontal pocket or fistula, occurring within an anatomical site of the jaw previously exposed to HN radiation. The focus is placed on vertical extent of necrosis on imaging and objective clinical findings of bone exposure or fistula formation and is named RadORN to reflect reliance on objective imaging findings. The length and duration of exposed bone, and subjective patient symptomatology were not considered a critical component of this system. The system recognizes 4 different Stages of ORN in addition to MBS (**Table 5**):

- *Stage-0* (sub-clinical, precursor of ORN): abnormal radiographic findings confined to alveolar bone only without clinical evidence. Patients with *Grade 0* ORN may or may not progress to overt disease.
- *Stage-1*: presence of exposed bone or bone probed through a periodontal pocket or fistula with any radiographic findings confined to alveolar bone.
- *Stage-2*: radiographic evidence of ORN progressed beyond alveolar bone and into basilar bone or sinus floor, in the absence of *Stage 3* findings.
- *Stage-3* (advanced ORN): *Stage 2* findings plus clinical and/or radiographic evidence of pathologic fracture, oro-antral or oro-nasal communication, and/or orocutaneous fistula.

**Table 5:**
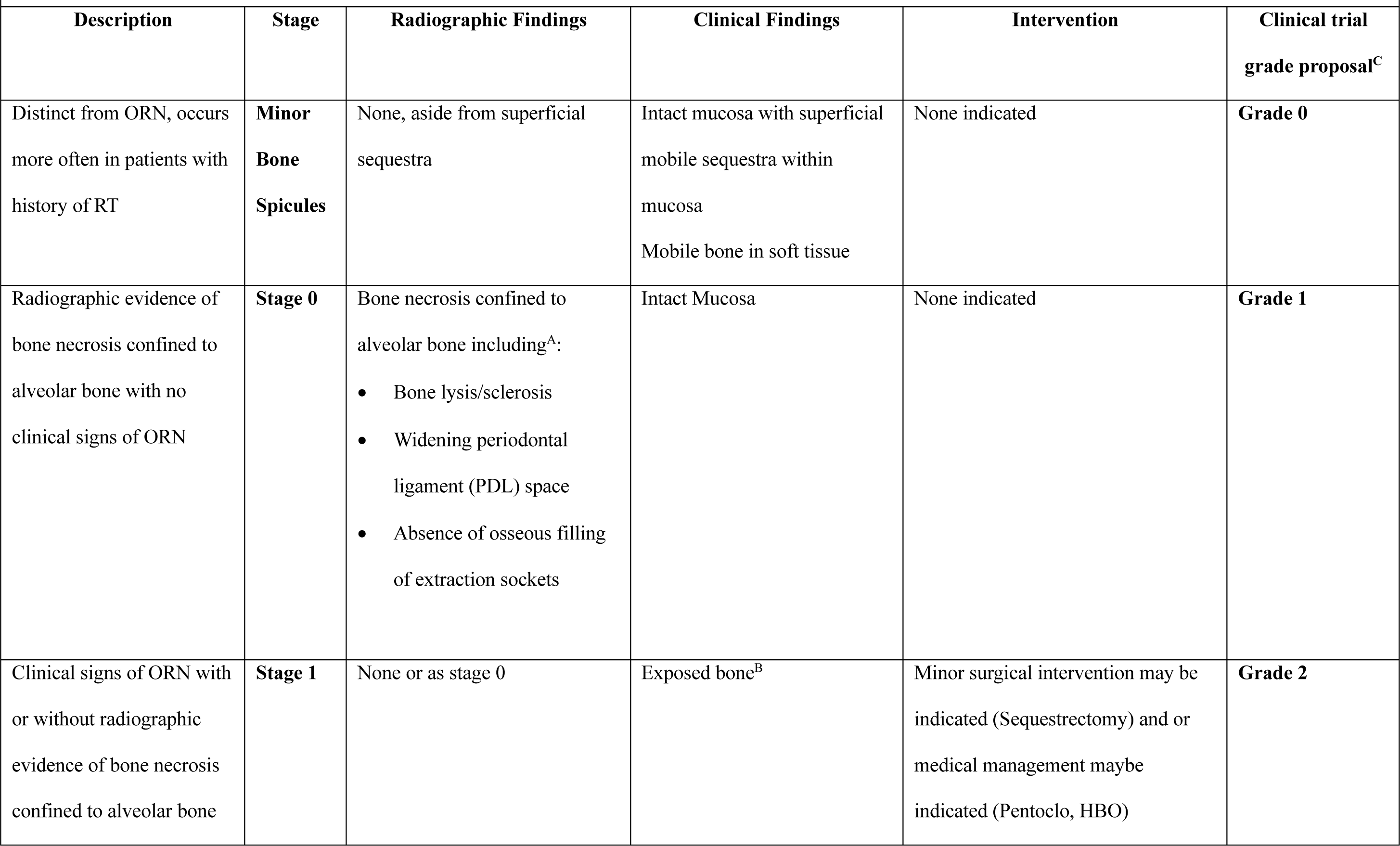
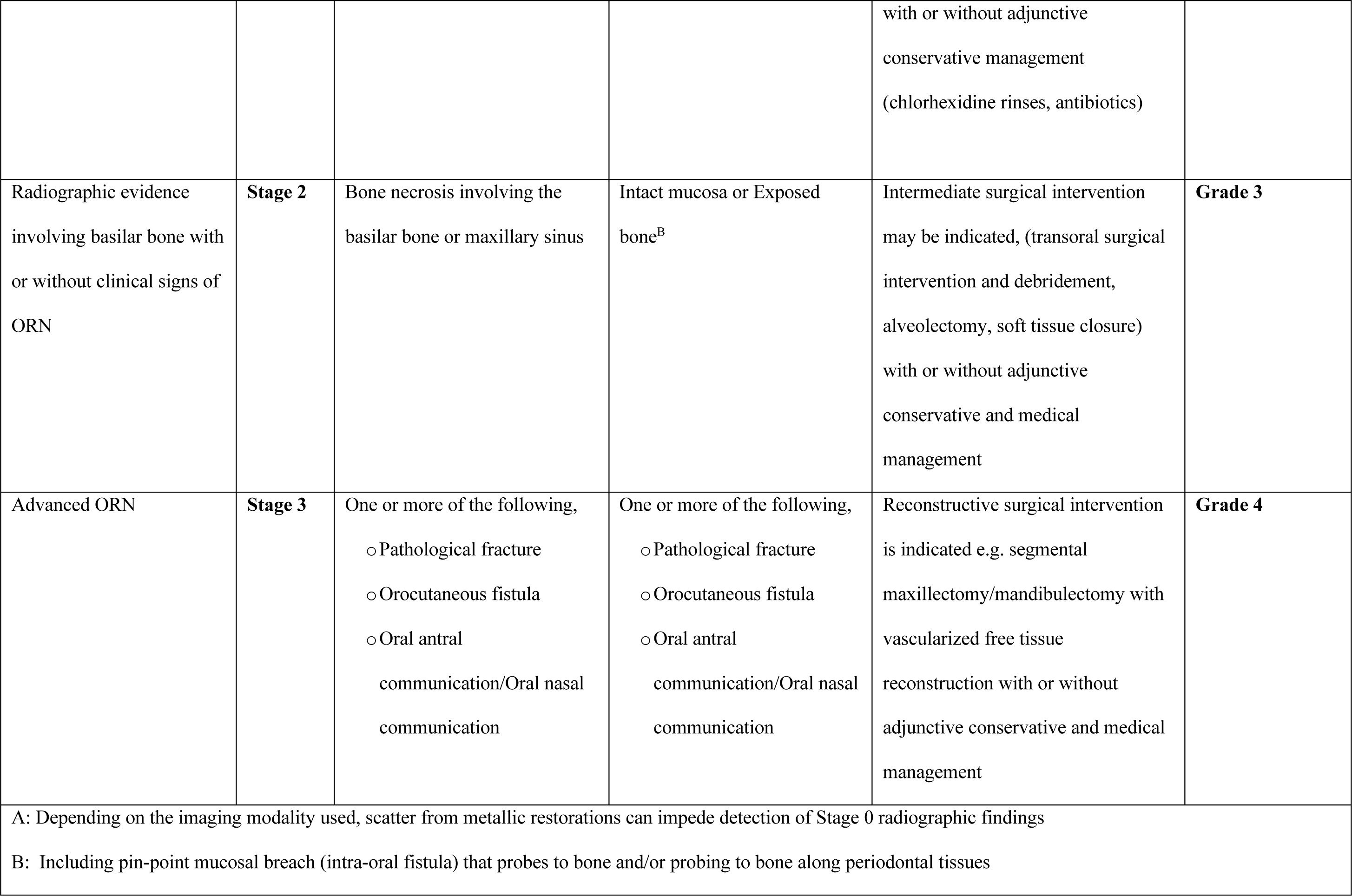
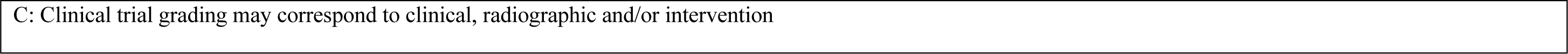
RadORN – A Novel Classification system for ORN.

Applying the RadORN system, we were able to classify all visits with ORN (0.0% missingness) and detected 5.7% of patients with the most severe stage of ORN. Patients with a more severe stage at time of ORN detection were more likely to develop serious ORN event (fracture or requiring surgical resection) at an earlier timepoint than patients with each successively less severe stage (**Figure 1**), with the caveat that stages 2 and 3 were grouped for statistical purpose due to low numbers of patients with stage 3 ORN at time of ORN detection. In addition, diagnosis of stage 2 ORN (HR 4.9, 95%CI 2.12-11.31, p<0.001) or stage 3 ORN (HR 11.45, 95%CI 3.34-39.32, p<0.001) significantly associated with risk of serious ORN event when compared to patients only diagnosed with the least severe stages. Results of time-dependent AUC analysis are presented in **Supplementary Table 2.** The RadORN system had numerically the highest c-index of all classification systems, though there was overlap in confidence intervals of the RadORN system versus other commonly-used systems such as Notani and Shaw. The RadORN system performed similarly, or better than, other grading systems when predicting serious ORN events at 1-, 2- and 3-years post-detection of ORN.

## DISCUSSION

This single institution study identifies the following patients with high-risk of developing ORN: OCC or OPC patients who received IMRT≥60Gy who are either current/ex-smoker and/or stage III-IV PC. A novel ORN classification system, RadORN, is proposed based on vertical extent of necrosis and/or bone exposure and fistula, which addressed limitations of existing systems and outperformed them in predicting serious ORN event. The RadORN system also harmonizes with the CTCAE classification to produce a grading system that can be used in clinical trials.

When considering the performance of previously proposed systems, the Notani classification system performed well across multiple domains. A major limitation of Notani is the inclusion of bone lysis beyond the inferior alveolar canal as stage 3 disease, as this means a patient with longstanding radiographic evidence of bone lysis beyond this landmark who then progress to fracture will not see an advancement in stage^18^. In fact, this characteristic resulted in the Notani classifying 13.2% of patients as having the most severe form of disease. In addition, Notani neglects to include the maxilla, a region where ORN is known to occur^30^. The Shaw system also performed well, which is predictable given that it is essentially the Notani system where some stage 1 cases are re-categorized as MBS^23^. The concept of MBS has been included is several previous classifications ^9,11^ and represents an entity distinct from ORN, known to occur in 0.2% of healthy patients following third molar extraction^31^. Incorporation of MBS into a classification system is important to prevent overdiagnosis of ORN, in particular for clinicians who maybe less familiar with this entity^23^. However, the Shaw classification did not perform better than Notani and had more missingness because it is more cumbersome with a requirement to measure size before applying the Notani system, making it impossible to generate a stage for each visit where size is not mentioned.

The CTCAE system performed well, as it characterizes ORN according to treatment need. However, the criteria for classification are subjective, and availability of treatment locally or local treatment paradigms would therefore influence severity of ORN. Grades 4, life-threatening consequences, and 5, Death due to ORN, were not observed to occur in this study. Despite this, CTCAE remains the preferred grading system for clinical trials, which present a unique challenge. Those recording outcomes may not be dental specialists who are accustomed to probing for exposed bone, limiting the detecting of stage I disease. Clinical trials also favour medical imaging as opposed to dental plain films. Where CT scans are used to assess response to therapy and monitor for recurrence, scatter from metallic restorations can significantly impede interpretation of regions surrounding teeth, making it difficult for medical imaging to diagnose Stage 0 disease. For these reasons, we harmonized our ORN classification system with the favoured clinical trials system, the CTCAE, to provide grading system that categories ORN as an adverse event based on commonly available clinical and medical imaging criteria, to be used in clinical trials (**Table 4**)

While the RTOG/EORTC and Schwartz systems performed well, these represent rudimentary measures of ORN based on bone response to radiation or vague radiographic findings that are difficult to apply clinically and had to be interpreted loosely for inclusion. The Store system also performed relatively well and while quite similar to the proposed system, we adopted radiographic only disease limited to alveolar bone as Stage-*0* and bone exposure with minimal radiographic findings as *Stage-1*, as does the AAOMS system for MRONJ, as opposed to the sequential ordering proposed by Store.

The AAOMS system for MRONJ was heavily consulted for the work, as this represents a system used almost universally to classify the disease. For this reason, there is excellent cross-study harmonization across studies on MRONJ, and the system is readily used in clinical practice to stage MRONJ in real-time. This makes the RadORN system familiar to dentists, in particular oral and maxillofacial surgeons. Given the numerous previously proposed classifications for ORN, we felt it important to incorporate characteristics favoured by both medical and dental professionals involved in management of ORN. Were a medical professional unfamiliar with the act of probing for exposed bone, ORN could still be staged according to common radiographic appearance, including subtle changes on dynamic contrast enhanced MRI, which at worst may understage stage I disease as stage 0 disease, where treatment recommendations do not significantly vary.

The inclusion of CTCAE in the treatment recommendations also seeks to align the system with current medical practice. In general, patients with Stage-0 disease correspond to CTCAE 1 – Patients who are asymptomatic with clinical or diagnostic observations only, where interventions are not indicated. Patients with Stage-1 ORN could correspond to CTCAE grades 1 or 2 depending on symptoms. These patients can generally be treated conservatively. Mobile sequestra may be removed with tooth preservation prioritized. Chlorhexidine may be recommended for prevention of infection in patients with exposed bone, while antibiotics may be prescribed for patients with signs and symptoms of active infection. Depending on the duration, size of the lesion, patient symptoms and availability of resources, some patients may be referred for medical management (PENTOCOL or HBO), or surgical management. Patients with Stage-2 ORN, with spread beyond alveolar bone, align with CTCAE grade 2 where medical or surgical interventions are indicated. Finally, Stage-3 ORN aligns with CTCAE grade 3 – severe symptoms with reconstructive surgical intervention is indicated, this being patients with pathologic fracture, orocutaneous fistula or oro-antral/nasal communication for which the gold standard for management remains segmental resection of disease segments and vascularized free tissue transfer ^32–34^.

Notably, the system does not include duration of exposure, size of bone exposure, symptoms or response to treatment. Analysis of systems that included a time or size component generally resulted in over-estimation of severity of ORN, higher missingness and non-significant performance on univariable analysis. While patient symptoms will dictate treatment, this is poorly correlated with extent of bone necrosis and we instead tied symptoms to management suggestions. Finally, response to therapy requires a patient to have completed a therapy before a stage can be generated, limiting its utility in treatment decision making and standardized data capture.

While the missingness for the RadORN system was zero, this is not surprising as 1) The system was designed using existing systems with low missingness, and 2) it was tested and adjusted using patient cases with high missingness across various systems. The RadORN system categorized 5.7% of patients with ORN as having the most severe form. This is similar to the proportion in systems that classify ORN in terms of functional deficits or required therapy, like CTCAE (6.6%). Systems that categorize spread of ORN beyond the inferior alveolar canal (Notani-13.2%, Shaw – 11.6%) appeared to over-estimate the proportion of patients with severe ORN, while simple binary systems or those with high missingness had artificially elevated results. When analyzing separability, Kaplan-Meier analysis demonstrated that the RadORN system performed as well as the Notani, Shaw, RTOG/EORTC and Store systems. Only Schwartz and Notani systems required no grouping of stages for statistical power. Given the high proportion of missingness for Karagozoglu and Schwartz, the results for these systems should be interpreted with caution. On univariable regression analysis, the proposed system outperformed other systems, demonstrating an appropriate stratification of disease from early to advanced ORN.

Our study has several limitations. The identified risk factors for development of ORN are based on a single-centre retrospective review. As expected, the RadORN system performed well when applied to data used to develop but has not been tested outside of this retrospective data set. Future studies will be needed to demonstrate the performance in a prospective clinical setting and datasets from outside of our institution. More detailed information about prescribed dose to structures at risk of ORN would provide greater relevance to the evaluation of risk for ORN. Results may not be replicable in an environment without reliable access to dental plain films.

## Conclusion

Our study identifies that oral-cavity or oropharyngeal primaries, received IMRT dose ≥60Gy, current/ex-smokers, and/or stage III-IV periodontal disease are ORN risk factors. We propose the RadORN system that classifies ORN based on vertical extent of bone necrosis and presence/absence of exposed bone/fistula: *Stage-0* (sub-clinical, precursor of ORN): abnormal radiographic findings confined to alveolar bone only without clinical evidence. *Stage-1*: the presence of exposed bone or bone probed through a periodontal pocket or fistula with any radiographic findings confined to alveolar bone. *Stage-2*: radiographic evidence of ORN beyond alveolar bone and into basilar bone or sinus floor. *Stage-3* (advanced ORN): *Stage-2* findings plus presence of jaw fracture or formation of fistula. The RadORN system outperformed existing systems in depicting the seriousness of ORN, and aligns with the CTCAE classification. It may facilitate clinical care and clinical trials.

## Data Availability

All data produced in the present study are available upon reasonable request to the corresponding author.

## Acknowledgements

Dr. Huang acknowledges support by the Bartley-Smith/Wharton, the Gordon Tozer, the Wharton Head and Neck Translational, Dr. Mariano Elia, the Joe’s Team, and the Petersen Funds at the Princess Margaret Foundation of author’s (SHH) academic activity. We acknowledge the efforts of the dental summer research students, Linnaea Halpert, Josh Moyal, Mariam Ghobrial, Ahuva Plonka and Lila Shapiro.

Dr. Fuller received/receives related funding and salary support from: NIH National Institute of Dental and Craniofacial Research (NIDCR) Establishing Outcome Measures for Clinical Studies of Oral and Craniofacial Diseases and Conditions award (R01DE025248), Exploratory/Developmental Research Grant Program (R21DE031082), and Prospective Observational or Biomarker Validation Study Cooperative Agreement Award (U01DE032168); a National Cancer Institute Parent Research Project Grant (R01CA258827).

## References

1. Singh A, Huryn JM, Kronstadt KL, Yom SK, Randazzo JR, Estilo CL. Osteoradionecrosis of the jaw: A mini review. Front Oral Health. 2022;3:980786. doi:10.3389/froh.2022.980786

2. Studer G, Bredell M, Studer S, Huber G, Glanzmann C. Risk profile for osteoradionecrosis of the mandible in the IMRT era. Strahlenther Onkol. Jan 2016;192(1):32–9. doi:10.1007/s00066-015-0875-6

3. Zhang W, Zhang X, Yang P, et al. Intensity-modulated proton therapy and osteoradionecrosis in oropharyngeal cancer. Radiother Oncol. Jun 2017;123(3):401–405. doi:10.1016/j.radonc.2017.05.006

4. Moon DH, Moon SH, Wang K, et al. Incidence of, and risk factors for, mandibular osteoradionecrosis in patients with oral cavity and oropharynx cancers. Oral Oncol. Sep 2017;72:98–103. doi:10.1016/j.oraloncology.2017.07.014

5. Ben-David MA, Diamante M, Radawski JD, et al. Lack of osteoradionecrosis of the mandible after intensity-modulated radiotherapy for head and neck cancer: likely contributions of both dental care and improved dose distributions. Int J Radiat Oncol Biol Phys. Jun 01 2007;68(2):396–402. doi:10.1016/j.ijrobp.2006.11.059

6. Wong K, Huang SH, O’Sullivan B, et al. Point-of-care outcome assessment in the cancer clinic: audit of data quality. Radiother Oncol. Jun 2010;95(3):339–43. doi:10.1016/j.radonc.2010.03.015

7. Watson E, Eason B, Kreher M, Glogauer M. The DMFS160: A new index for measuring post-radiation caries. Oral Oncol. Sep 2020;108:104823. doi:10.1016/j.oraloncology.2020.104823

8. Caton JG, Armitage G, Berglundh T, et al. A new classification scheme for periodontal and peri-implant diseases and conditions - Introduction and key changes from the 1999 classification. J Periodontol. Jun 2018;89 Suppl 1:S1–S8. doi:10.1002/JPER.18-0157

9. Coffin F. The incidence and management of osteoradionecrosis of the jaws following head and neck radiotherapy. Br J Radiol. Nov 1983;56(671):851–7. doi:10.1259/0007-1285-56-671-851

10. Marx RE, Johnson RP. Studies in the radiobiology of osteoradionecrosis and their clinical significance. Oral Surg Oral Med Oral Pathol. Oct 1987;64(4):379–90. doi:10.1016/0030-4220(87)90136-8

11. Morton ME, Simpson W. The management of osteoradionecrosis of the jaws. Br J Oral Maxillofac Surg. Oct 1986;24(5):332–41. doi:10.1016/0266-4356(86)90018-5

12. Cox JD, Stetz J, Pajak TF. Toxicity criteria of the Radiation Therapy Oncology Group (RTOG) and the European Organization for Research and Treatment of Cancer (EORTC). Int J Radiat Oncol Biol Phys. Mar 30 1995;31(5):1341–6. doi:10.1016/0360-3016(95)00060-C

13. Glanzmann C, Grätz KW. Radionecrosis of the mandibula: a retrospective analysis of the incidence and risk factors. Radiother Oncol. Aug 1995;36(2):94–100. doi:10.1016/0167-8140(95)01583-3

14. Clayman L. Clinical controversies in oral and maxillofacial surgery: Part two. Management of dental extractions in irradiated jaws: a protocol without hyperbaric oxygen therapy. J Oral Maxillofac Surg. Mar 1997;55(3):275–81. doi:10.1016/s0278-2391(97)90542-5

15. LENT SOMA tables. Radiother Oncol. Apr 1995;35(1):17–60.

16. Støre G, Boysen M. Mandibular osteoradionecrosis: clinical behaviour and diagnostic aspects. Clin Otolaryngol Allied Sci. Oct 2000;25(5):378–84. doi:10.1046/j.1365-2273.2000.00367.x

17. Schwartz HC, Kagan AR. Osteoradionecrosis of the mandible: scientific basis for clinical staging. Am J Clin Oncol. Apr 2002;25(2):168–71. doi:10.1097/00000421-200204000-00013

18. Notani K, Yamazaki Y, Kitada H, et al. Management of mandibular osteoradionecrosis corresponding to the severity of osteoradionecrosis and the method of radiotherapy. Head Neck. Mar 2003;25(3):181–6. doi:10.1002/hed.10171

19. Tsai CJ, Hofstede TM, Sturgis EM, et al. Osteoradionecrosis and radiation dose to the mandible in patients with oropharyngeal cancer. Int J Radiat Oncol Biol Phys. Feb 01 2013;85(2):415–20. doi:10.1016/j.ijrobp.2012.05.032

20. Lyons A, Osher J, Warner E, Kumar R, Brennan PA. Osteoradionecrosis--a review of current concepts in defining the extent of the disease and a new classification proposal. Br J Oral Maxillofac Surg. May 2014;52(5):392–5. doi:10.1016/j.bjoms.2014.02.017

21. Common Terminology Criteria for Adverse Events (CTCAE) Version 5 (Nov 27, 2017).

22. Caparrotti F, Huang SH, Lu L, et al. Osteoradionecrosis of the mandible in patients with oropharyngeal carcinoma treated with intensity-modulated radiotherapy. Cancer. Oct 01 2017;123(19):3691–3700. doi:10.1002/cncr.30803

23. Shaw R, Tesfaye B, Bickerstaff M, Silcocks P, Butterworth C. Refining the definition of mandibular osteoradionecrosis in clinical trials: The cancer research UK HOPON trial (Hyperbaric Oxygen for the Prevention of Osteoradionecrosis). Oral Oncol. Jan 2017;64:73–77. doi:10.1016/j.oraloncology.2016.12.002

24. Ruopp MD, Perkins NJ, Whitcomb BW, Schisterman EF. Youden Index and optimal cut-point estimated from observations affected by a lower limit of detection. Biom J. Jun 2008;50(3):419–30. doi:10.1002/bimj.200710415

25. Bansal A, Heagerty PJ. A Tutorial on Evaluating the Time-Varying Discrimination Accuracy of Survival Models Used in Dynamic Decision Making. Med Decis Making. Nov 2018;38(8):904–916. doi:10.1177/0272989X18801312

26. Kamarudin AN, Cox T, Kolamunnage-Dona R. Time-dependent ROC curve analysis in medical research: current methods and applications. BMC Med Res Methodol. Apr 07 2017;17(1):53. doi:10.1186/s12874-017-0332-6

27. Marx RE. A new concept in the treatment of osteoradionecrosis. J Oral Maxillofac Surg. Jun 1983;41(6):351–7. doi:10.1016/s0278-2391(83)80005-6

28. Ruggiero SL, Dodson TB, Fantasia J, et al. American Association of Oral and Maxillofacial Surgeons position paper on medication-related osteonecrosis of the jaw--2014 update. J Oral Maxillofac Surg. Oct 2014;72(10):1938–56. doi:10.1016/j.joms.2014.04.031

29. Ruggiero SL, Dodson TB, Aghaloo T, Carlson ER, Ward BB, Kademani D. American Association of Oral and Maxillofacial Surgeons’ Position Paper on Medication-Related Osteonecrosis of the Jaws-2022 Update. J Oral Maxillofac Surg. May 2022;80(5):920–943. doi:10.1016/j.joms.2022.02.008

30. Shokri T, Wang W, Vincent A, Cohn JE, Kadakia S, Ducic Y. Osteoradionecrosis of the Maxilla: Conservative Management and Reconstructive Considerations. Semin Plast Surg. May 2020;34(2):106–113. doi:10.1055/s-0040-1709144

31. Bui CH, Seldin EB, Dodson TB. Types, frequencies, and risk factors for complications after third molar extraction. J Oral Maxillofac Surg. Dec 2003;61(12):1379–89. doi:10.1016/j.joms.2003.04.001

32. Baumann DP, Yu P, Hanasono MM, Skoracki RJ. Free flap reconstruction of osteoradionecrosis of the mandible: a 10-year review and defect classification. Head Neck. Jun 2011;33(6):800–7. doi:10.1002/hed.21537

33. Chang DW, Oh HK, Robb GL, Miller MJ. Management of advanced mandibular osteoradionecrosis with free flap reconstruction. Head Neck. Oct 2001;23(10):830–5. doi:10.1002/hed.1121

34. O’Connell JE, Brown JS, Rogers SN, Bekiroglu F, Schache A, Shaw RJ. Outcomes of microvascular composite reconstruction for mandibular osteoradionecrosis. Br J Oral Maxillofac Surg. Nov 2021;59(9):1031–1035. doi:10.1016/j.bjoms.2020.11.013

