## Supplementary Table 2 for "Development and Standardization of a Classification System for Osteoradionecrosis: Implementation of a Risk-Based Model"

### Time-dependent Area-under-curve (AUC) C-statistic

- C-statistic – measures discrimination between patients with adverse events vs no event across the entire time-to-event curve

#### I/D Time-dependent AUC comparison, all graded visits

C-index and I/D Time-Dependent AUC, All Graded Visits

|  | C index (95% CI) | AUC, 1 year (95% CI) | AUC, 2 years (95% CI) | AUC, 3 years (95% CI) |
| --- | --- | --- | --- | --- |
| Proposed.system | 0.52 (0.31,0.61) | 0.55 (0.39, 0.74) | 0.62 (0.46, 0.74) | 0.34 (0.25, 0.61) |
| Notani | 0.49 (0.31,0.59) | 0.41 (0.27, 0.59) | 0.59 (0.43, 0.74) | 0.41 (0.30, 0.71) |
| STORE | 0.48 (0.27,0.59) | 0.47 (0.33, 0.68) | 0.58 (0.41, 0.73) | 0.44 (0.24, 0.69) |
| PMCC | 0.45 (0.29,0.56) | 0.54 (0.35, 0.70) | 0.44 (0.23, 0.60) | 0.55 (0.37, 0.74) |
| SCHWARTZ | 0.42 (0.25,0.52) | 0.35 (0.21, 0.58) | 0.54 (0.35, 0.71) | 0.37 (0.22, 0.60) |
| KARAGOZOGLU | 0.42 (0.23,0.52) | 0.36 (0.20, 0.54) | 0.54 (0.32, 0.71) | 0.49 (0.28, 0.70) |
| Shaw | 0.41 (0.25,0.55) | 0.46 (0.20, 0.70) | 0.61 (0.34, 0.81) | 0.45 (0.26, 0.76) |
| SOMA | 0.37 (0.23,0.51) | 0.34 (0.21, 0.51) | 0.32 (0.16, 0.53) | 0.44 (0.23, 0.65) |
| RTOG | 0.36 (0.24,0.49) | 0.43 (0.30, 0.62) | 0.55 (0.34, 0.70) | 0.31 (0.15, 0.57) |
| TSAI | 0.34 (0.20,0.48) | 0.39 (0.20, 0.60) | 0.32 (0.11, 0.59) | 0.46 (0.23, 0.71) |
| CTCAE | 0.32 (0.20,0.44) | 0.25 (0.16, 0.42) | 0.42 (0.21, 0.60) | 0.37 (0.17, 0.62) |

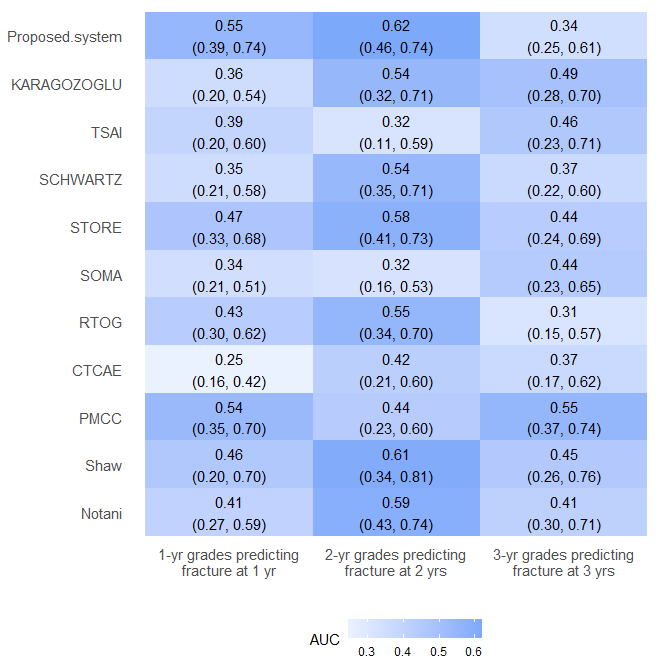

Difference in ID AUCs between Proposed System versus other grading systems (with bootstrapped 95% CIs)

| System | Difference, 1 year | Difference, 2 years | Difference, 3 years |
| --- | --- | --- | --- |
| Notani | 0.14 (-0.11,0.36) | 0.03 (-0.20,0.23) | -0.07 (-0.35,0.20) |
| Shaw | 0.09 (-0.21,0.38) | 0.01 (-0.24,0.28) | -0.11 (-0.39,0.24) |
| PMCC | 0.01 (-0.20,0.28) | 0.18 (-0.03,0.42) | -0.21 (-0.41,0.12) |
| CTCAE | 0.30 (0.07,0.49) | 0.20 (-0.03,0.44) | -0.03 (-0.28,0.31) |
| RTOG | 0.11 (-0.15,0.34) | 0.07 (-0.17,0.32) | 0.03 (-0.21,0.33) |
| SOMA | 0.20 (-0.03,0.42) | 0.30 (0.02,0.48) | -0.10 (-0.32,0.25) |
| STORE | 0.08 (-0.17,0.28) | 0.04 (-0.19,0.26) | -0.09 (-0.34,0.23) |
| SCHWARTZ | 0.20 (-0.07,0.42) | 0.08 (-0.15,0.30) | -0.03 (-0.26,0.25) |
| TSAI | 0.15 (-0.14,0.41) | 0.30 (-0.03,0.53) | -0.12 (-0.35,0.24) |
| KARAGOZOGLU | 0.19 (-0.06,0.43) | 0.08 (-0.15,0.31) | -0.15 (-0.35,0.21) |
| Proposed.system | - | - | - |

C/D Time-Dependent AUC, All Graded Visits

|  | AUC, 1 year (95% CI) | AUC, 2 years (95% CI) |
| --- | --- | --- |
| Proposed.system | 0.58 (0.41, 0.81) | 0.71 (0.48, 0.86) |
| KARAGOZOGLU | 0.72 (0.54, 0.86) | 0.69 (0.41, 0.94) |
| TSAI | 0.60 (0.39, 0.81) | 0.46 (0.19, 0.78) |
| SCHWARTZ | 0.75 (0.66, 0.85) | 0.65 (0.32, 0.92) |
| STORE | 0.78 (0.64, 0.88) | 0.64 (0.31, 0.89) |
| SOMA | 0.64 (0.44, 0.82) | 0.46 (0.23, 0.70) |
| RTOG | 0.56 (0.36, 0.76) | 0.58 (0.32, 0.86) |
| CTCAE | 0.49 (0.29, 0.68) | 0.57 (0.30, 0.83) |
| PMCC | 0.60 (0.36, 0.81) | 0.71 (0.49, 0.90) |
| Shaw | 0.73 (0.52, 0.91) | 0.66 (0.34, 0.98) |
| Notani | 0.78 (0.70, 0.87) | 0.70 (0.44, 0.93) |

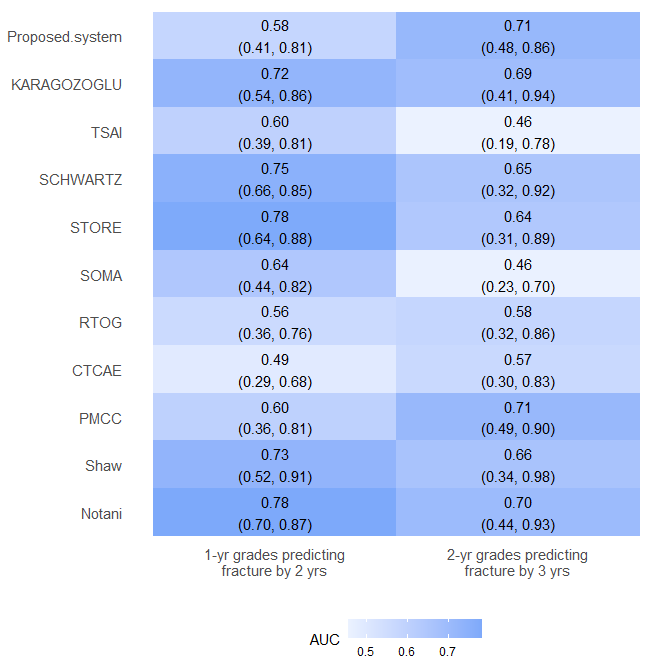

Difference in ID AUCs between Proposed System versus other grading systems (with bootstrapped 95% CIs)

| System | Difference, 1-yr grades | Difference, 2-yr grades |
| --- | --- | --- |
| Notani | -0.20 (-0.40,0.03) | 0.01 (-0.34,0.34) |
| Shaw | -0.15 (-0.41,0.15) | 0.05 (-0.33,0.42) |
| PMCC | -0.02 (-0.30,0.29) | 0.00 (-0.28,0.28) |
| CTCAE | 0.09 (-0.17,0.38) | 0.15 (-0.22,0.46) |
| RTOG | 0.02 (-0.24,0.32) | 0.14 (-0.21,0.44) |
| SOMA | -0.07 (-0.33,0.23) | 0.25 (-0.06,0.54) |
| STORE | -0.20 (-0.40,0.05) | 0.07 (-0.29,0.44) |
| SCHWARTZ | -0.17 (-0.37,0.08) | 0.06 (-0.29,0.41) |
| TSAI | -0.02 (-0.32,0.26) | 0.26 (-0.14,0.56) |
| KARAGOZOGLU | -0.15 (-0.38,0.14) | 0.03 (-0.32,0.36) |
| Proposed.system | - | - |
